# Quality indicators on medicines packaging as guide to routine quality assurance for procured generic medicines: A community pharmacy survey in Nigeria

**DOI:** 10.1101/2022.10.22.22281401

**Authors:** Ebiowei S.F Orubu, Elamene C.O. Ukuta, Faith O. Robert

## Abstract

**Introduction:** While medicines quality assurance is best performed by a Stringent Regulatory Authority (SRA), many countries still lack this capacity. In these contexts, brand manufacturers take additional steps to signal authenticity to their consumers through the use of overt authentication technologies on medicines packaging. The aim of this study was to use quality indicators on the secondary packaging of medicines to identify, and, thus, risk rank, therapeutic groups most prone to being substandard or falsified.

**Method:** This study profiled medicines in the inventory of a conveniently selected pharmacy in Nigeria by three common packaging quality indicators: (i) National Agency for Food and Drug Administration, NAFDAC, registration number, (ii) a mobile authentication service, MAS, number, and (iii) a hologram.

**Results:** The inventory contained 66 medicines available as 380 oral, parenteral, or topical products. Of these, 92% had a NAFDAC number, 20% had a MAS number, and 10% bore a hologram – either exclusively, or simultaneously. Antimicrobials – mostly antibiotics and antimalarials – were the most common therapeutic group containing one or all three quality indications. Antibiotic medicines for children formulated as suspensions and drops had the most quality assurance indicators on their packaging.

**Conclusion:** This result suggests that antimicrobials, especially those formulated for use in children, may be the medicine group at the greatest risk of being substandard or falsified (SF) in Nigeria. **The implication of this finding is the need for healthcare facilities to implement strict medicines quality assurance procurement practices especially for antimicrobials**. Continued vigilance by the public would also be necessary.

## Introduction

Substandard and falsified (SF) medicines are a top 10 global public health challenge. The global prevalence of SF medicines is estimated at 10%, with the greater incidence in in Asia and Africa (1). SF medicines negatively impacts the health and wealth of nations (2,3). This problem compounds existing socioeconomic and infrastructural challenges predisposing to poor health outcomes and calls for urgent solutions to ensure public health.

### Regulatory system capacity strengthening is the preferred option for ensuring the integrity of the supply chain

Medicine quality assurance systems rests upon a Stringent Regulatory Authority (SRA) with the legal backing and requisite resources to ensure protection of the pharmaceutical supply chain against the entry of SF medicines. This service involves multiple steps in the pathway from manufacturing to dispensing or use at the patient level. However, this regulatory capacity is lacking in many Low- and Middle-Income Countries (LMICs). In Africa, only Egypt, Ghana, Nigeria and Tanzania meet the World Health Organization (WHO)’s Global Benchmarking Criteria for qualification as a SRA, or a regulatory authority operating at Maturity Level 3 or 4, that is, with the capacity to continuously ensure the quality of medicines in circulation (4).

Involving other stakeholders, including, consumers in post-marketing medicines quality surveillance could help reduce the incidence of SF medicines. In the early 2000s’, Nigeria launched one such program. “Operation Shine Your Eyes” was a public awareness program ran on mass media by the National Agency for Food and Drug Administration (NAFDAC), the national medicines regulatory authority, aimed at increasing awareness. This program alerted consumers on the need to look for a NAFDAC’s number as a sign of quality. This intervention and others instituted by NAFDAC is estimated to have reduced the prevalence of SF medicines, dropping over the five-year period of 2001 to 2006 from 41% to 16.7% (5).

Subsequently, manufacturers stepped up this initiative of involving consumers in medicines quality assurance with the launch of several authentication technologies, or Mobile Authentication Services (MAS) including short codes revealed after removing a foil placed on the outer or secondary package of the medicine. This technology leveraged the use of mobile phones (and the similarity of the action of using mobile phone top-up processes). In 2012, NAFDAC mandated the MAS across antibiotics and antimalarials (6).

Public perception of a high prevalence of poor quality medicines persist in Nigeria, despite the efforts of NAFDAC (7–10). Medicines procurement practices do not routinely include quality assurance in Nigeria. There is the need for public healthcare facilities to integrate routine medicines quality assurance into their procurement practices.

The aim of this study was to identify medicine groups most at risk of being substandard or falsified in Nigeria to guide routine quality assurance during procurement. The study assumed that medicines groups most likely to be SF would bear the most quality indicators on their packaging, and, thus, predict risk of SF medicines among generics.

## Method

### Study design

The study was designed as a case study using a purposively selected and conveniently located community pharmacy in Nigeria.

### Study location and setting

Study location and setting were both conveniently selected based on access to data. Study location was Bayelsa State in the South-South geopolitical zone of Nigeria (Figure 1). Study setting was a community pharmacy. The pharmacy is run professionally, with a superintendent pharmacist overseeing other front-facing staff who are trained pharmacy technicians.

**Figure 1.**
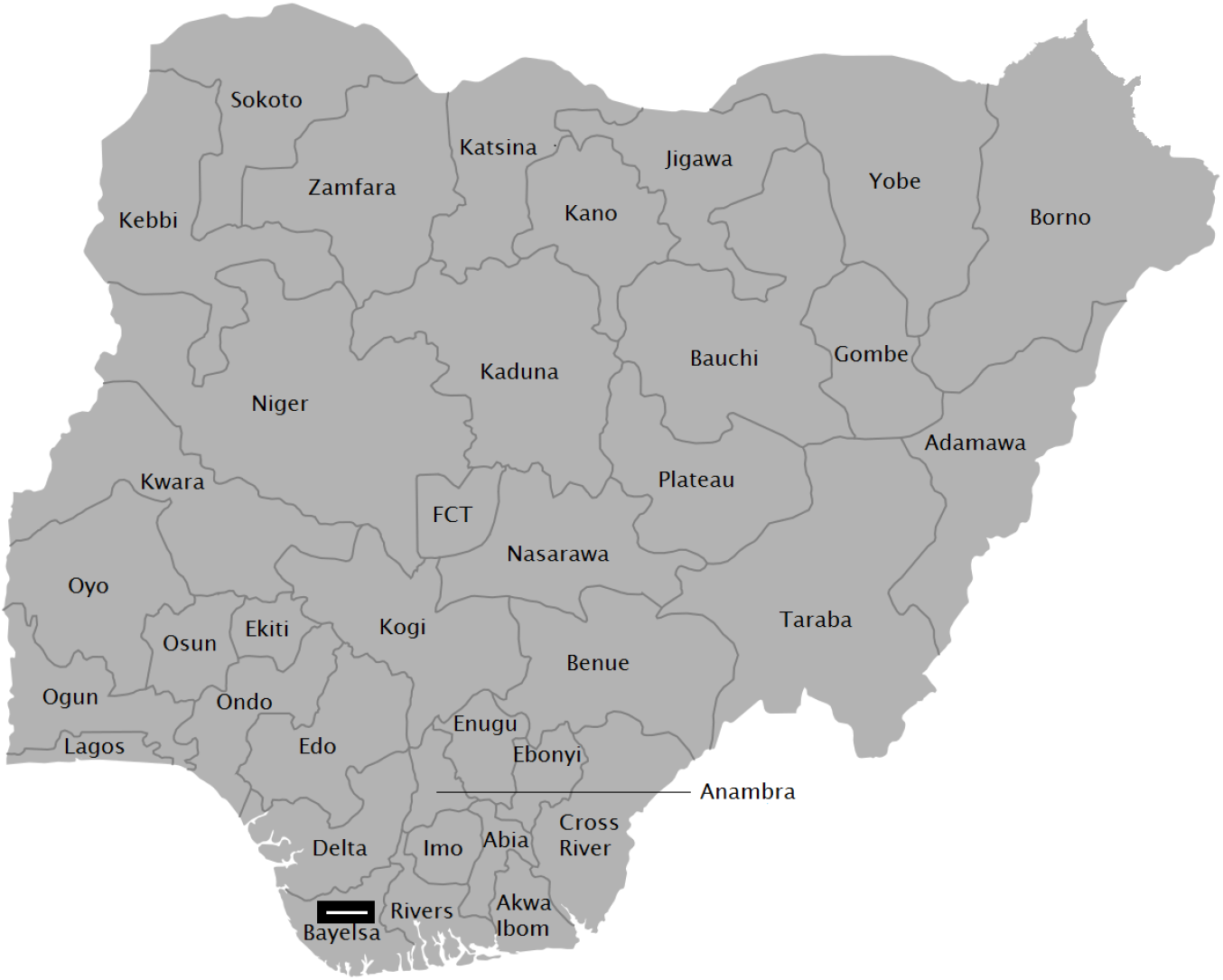
Geographical location of the study site indicated by the rectangle on the map. [Source: Modified from: https://en.wikipedia.org/wiki/File:Map_of_Nigerian_States_with_names.png#/media/File:Map_of_Nigerian_States_with_names.png]

### Sample

All medicines held in stock as at April 30 2018 were included in the study. Data extracted from samples were: INN, brand name, formulation (commercial product comprising a distinct combination of dosage form, whether tablet, capsule, injection, syrup, or eyedrop for example, and dosage form strength; for example Ampiclox capsules 500mg), manufacturer, country of origin of the manufacturer, batch number, dates of manufacture and expiry, pack size, presence or absence of NAFDAC number and the number itself when present, and two other authentication technologies – hologram or scratch code. These two authentication technologies were selected as, from experience, they were the more common authentication technologies in the Nigerian market as well as being simpler to recognize.

### Data analysis

The WHO/ATC system was used to describe and analyze collected data by descriptive statistics. Product characteristics were expressed in percentages, with inventory composition described at the anatomic, level 1, of the WHO/ATC classification. Products were also characterized in terms of source, whether as imports or locally-produced. Imports were noted by country. The occurrence of the three selected quality indicators were described for the entire inventory at the anatomic main group level. Formulations with the greatest number of quality indicators – all three selected – were noted in terms of indications for use in adults or children.

## Results

### Product characteristics

The inventory consisted of 66 unique INNs available as 380 formulations or commercial products. Oral formulations were the most common, 59% (226/380); followed by parenteral products, 24% (92/360), and topicals, 17% (62/380).

The products covered 12 of the 14 anatomical/therapeutic groups in the ATC classification, but for Antineoplastics and Immunomodulating Agents (Table 1). Anti-infectives for systemic use, A01, was the dominant therapeutic group at the WHO/ATC level I classification, comprising about a third (28%, 108/380) of all products, followed by Alimentary Track and Metabolism (12%, 45/380), Nervous System and Cardiovascular System, each at 9% (respectively, 35/380 and 33/380). Antibacterials for systemic use, J01, were the most prominent level 2 products, comprising 27% (102/380) of products.

**Table 1.**
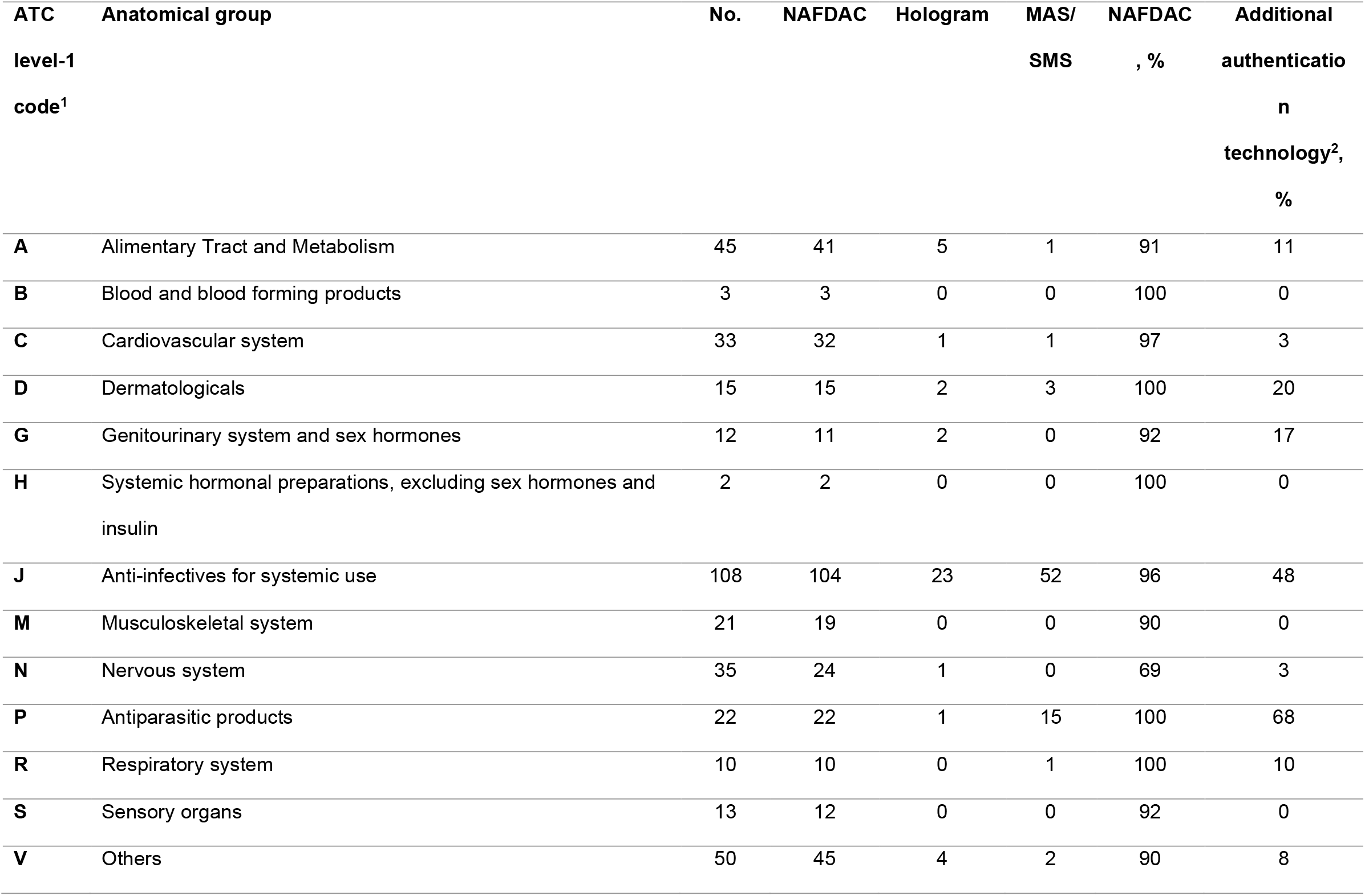
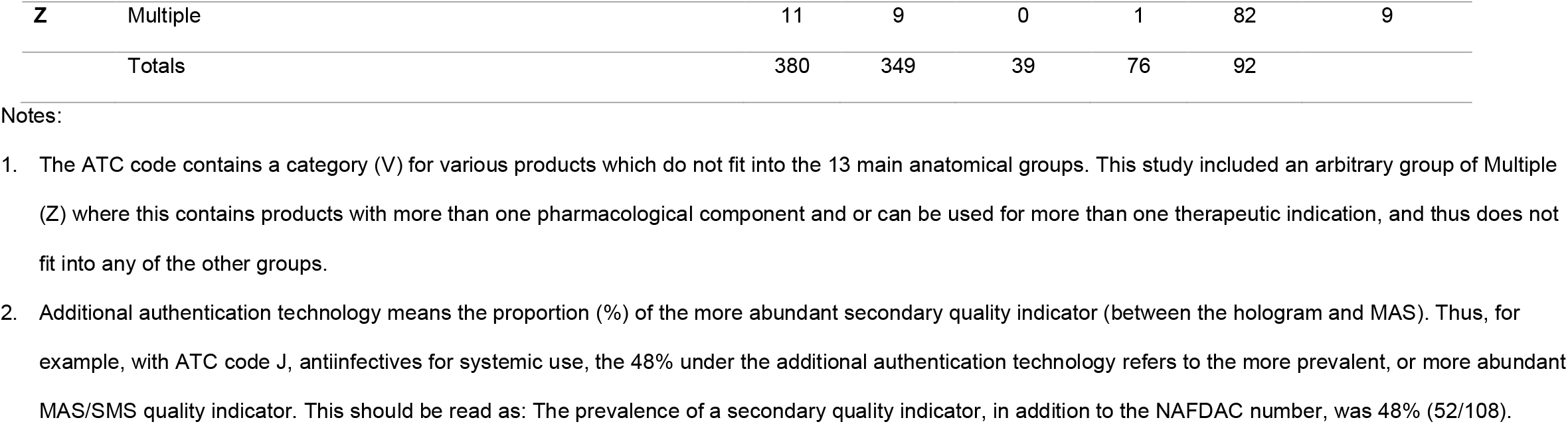
Prevalence of quality indicators by anatomical group for a sample of 380 commercial products at a pharmacy in Bayelsa State Nigeria

| **ATC level-1 code<sup>1</sup>** | **Anatomical group** | **No.** | **NAFDAC** | **Hologram** | **MAS/ SMS** | **NAFDAC , %** | **Additional authentication technology<sup>2</sup>, %** |
| --- | --- | --- | --- | --- | --- | --- | --- |
| **A** | Alimentary Tract and Metabolism | 45 | 41 | 5 | 1 | 91 | 11 |
| **B** | Blood and blood forming products | 3 | 3 | 0 | 0 | 100 | 0 |
| **C** | Cardiovascular system | 33 | 32 | 1 | 1 | 97 | 3 |
| **D** | Dermatologicals | 15 | 15 | 2 | 3 | 100 | 20 |
| **G** | Genitourinary system and sex hormones | 12 | 11 | 2 | 0 | 92 | 17 |
| **H** | Systemic hormonal preparations, excluding sex hormones and insulin | 2 | 2 | 0 | 0 | 100 | 0 |
| **J** | Anti-infectives for systemic use | 108 | 104 | 23 | 52 | 96 | 48 |
| **M** | Musculoskeletal system | 21 | 19 | 0 | 0 | 90 | 0 |
| **N** | Nervous system | 35 | 24 | 1 | 0 | 69 | 3 |
| **P** | Antiparasitic products | 22 | 22 | 1 | 15 | 100 | 68 |
| **R** | Respiratory system | 10 | 10 | 0 | 1 | 100 | 10 |
| **S** | Sensory organs | 13 | 12 | 0 | 0 | 92 | 0 |
| **V** | Others | 50 | 45 | 4 | 2 | 90 | 8 |
| **Z** | Multiple | 11 | 9 | 0 | 1 | 82 | 9 |
|  | Totals | 380 | 349 | 39 | 76 | 92 |  |

Notes:
1. The ATC code contains a category (V) for various products which do not fit into the 13 main anatomical groups. This study included an arbitrary group of Multiple (Z) where this contains products with more than one pharmacological component and or can be used for more than one therapeutic indication, and thus does not fit into any of the other groups. 2. Additional authentication technology means the proportion (%) of the more abundant secondary quality indicator (between the hologram and MAS). Thus, for example, with ATC code J, antiinfectives for systemic use, the 48% under the additional authentication technology refers to the more prevalent, or more abundant MAS/SMS quality indicator. This should be read as: The prevalence of a secondary quality indicator, in addition to the NAFDAC number, was 48% (52/108).

Antimicrobial products – antibiotics, antimalarials, anti-parasitics, antivirals, antimycotics, anthelminthics, antifungals, and vaccines – comprising ATC level 1 J and P groups, comprised almost half: 44% (167/380) of the inventory. Antimicrobials were dominated by six therapeutic and pharmacological sub-groups constituting the majority 75%: penicillins (24%, 40/165); cephalosporins (15%, 25/165); antimycotics, or antifungals, (14%, 23/165); antimalarials (10%, 17/165); fluroquinolones (6%, 10/165); and macrolides (6%, 10/165).

Imports were the greater source of medicines (79%, 299/380). Almost all imports, by geographical location, were either from Asia (59%, 175/299), or Europe, including the United Kingdom, (36%, 107/299). However, by specific countries, the largest source of medicines was India (31%, 116/380), followed by Nigeria (21%, 81/380).

### Quality indicators

The majority (92%, 351/380) of formulations had a NAFDAC number (Figure 2).

**Figure 2.**
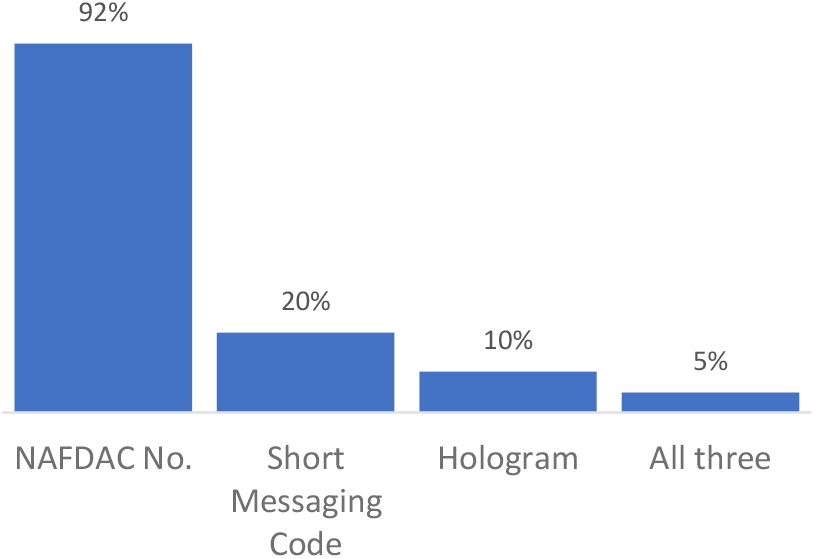
Prevalence of medicines quality indicators on the packaging of 380 medicines available at a community pharmacy in Bayelsa State, Nigeria. Prevalence was not mutually exclusive. The presence of a NAFDAC number, for example, did not rule out the presence of another quality indicator. Hence the sum of all quality indicators was greater than 100%.

Only 10% (39/380) of the products had a hologram – regardless of the presence or absence of a NAFDAC number (Figure 2). Holograms were found mostly (67%, 26/39) on antimicrobials – antibacterials and antiparasitics; and occur almost exclusively (97%, 38/39) on imports.

The incidence of MAS was 20% (76/380) (Figure 2). The majority (96%, 73/76) of products with MAS were also antimicrobials. Only 3 formulations out of 76 with scratch codes were non-antimicrobials – salbutamol inhaler, a multivitamin product, Pregnacare™, and a calcium channel blocker, amlodipine. Products with MAS were also mostly imports (92%, 70/76).

The prevalence of all three authentication technologies was 5% (17/380). These 17 products were all antibiotics (beta-lactams, quinolones, and macrolides). All were imports (Table 2). These products were mostly (59%, 10/17) penicillins – amoxicillin, ampicillin-cloxacillin, and amoxicillin-clavulanate (Table 2).

**Table 2.** Characteristics of products with all three medicines quality indicators on packages in a selected community pharmacy in Nigeria, 2018

| Product<br>NAME | GENERIC NAME | TYPE | STRENGTH | PACK<br>SIZE | DOSAGE<br>FORM | ATC<br>CODE | HOLOGRAM<br>OR SEAL | MAS | NAFDAC<br>NO. | ORIGIN |
| --- | --- | --- | --- | --- | --- | --- | --- | --- | --- | --- |
| **Amoxil** | Amoxicillin | Brand | 125mg/5ml | 70mls | Suspension | J01C | YES | YES | 04 2402 | INDIA |
| **Augmentin** | Amoxicillin+Clavulanate | Brand | 625mg | 2x7 | Tablet | J01C | YES | YES | 04 1928 | U. K |
| **Augmentin** | Amoxicillin+Clavulanate | Brand | 1g | 2x7 | Tablet | J01C | YES | YES | 04 8655 | U. K |
| **Augmentin** | Amoxicillin+Clavulanate | Brand | 228mg/5ml | 70mls | Suspension | J01C | YES | YES | 04 2516 | U. K |
| **Amoksiklav** | Amoxicillin+Clavulanic<br>Acid | Branded Generic | 457/5ml | 70mls | Suspension | J01C | YES | YES | A4 0882 | SLOVENIA |
| **Amoksiklav** | Amoxicillin+Clavulanic<br>Acid | Brand | 312.5mg/5ml | 100ml | Suspension | J01C | YES | YES | 04 2732 | SLOVENIA |
| **Amoksiklav** | Amoxicillin+Clavulanic<br>Acid | Brand | 156.25mg/5ml | 100ml | Suspension | J01C | YES | YES | 04 2626 | SLOVENIA |
| **Amovin** | Amoxicillin+Clavulanic<br>Acid | Brand | 228.5mg/5ml | 100ml | Suspension | J01C | YES | YES | 04 8405 | INDIA |
| **Ampiclox** | Ampicillin+Cloxacillin | Brand | 250mg/5ml | 100mls | Suspension | J01C | YES | YES | 04 2580 | INDIA |
| **Ampiclox** | Ampicillin+Cloxacillin | Brand | 90mg/0.6ml | 8mls | Drop | J01C | YES | YES | 04 3576 | INDIA |
| **Suitrox** | Azithromycin | Branded Generic | 200mg/5ml | 20ml | Suspension | J01F | YES | YES | A4 3463 | ROMANIA |
| **Rocephine** | Ceftriaxone | Brand | 1g | 1 | Injection | J01D | YES | YES | 04 7302 | SWITZERLAND |
| **Zinnat** | Cefuroxime | Brand | 500mg | 1×10 | Tablet | J01D | YES | YES | A4 4880 | U. K |
| **Zinnat** | Cefuroxime | Brand | 125mg/5ml | 100ml | Suspension | J01D | YES | YES | 04 0465 | U. K |
| **Sivoryl** | Erythromycin | Branded Generic | 500mg | 1×10 | Tablet | J01F | YES | YES | B4 1844 | INDIA |
| **Jozza** | Ofloxacin | Branded Generic | 400mg | 1×10 | Tablet | J01M | YES | YES | B4 7048 | INDIA |
| **Peflomed** | Pefloxacin | Branded Generic | 400mg | 2×7 | Tablet | J01M | YES | YES | 04 3153 | INDIA |

Notes: 1. Products listed at the ATC level 3, or pharmacological sub-group level. 2. JO1C = Beta-lactam antibiotics, penicillins; JO1D = Beta-lactam antibiotics, others; JO1F = Macrolides, lincosamides, streptogramins; JO1M = Quinolones. 3. MAS = Mobile Authentication Service (short code). 4. NAFDAC = National Agency for Drug Administration and Control.

Most of these (59%, 10/17) were antibiotic medicines formulated for use in children as suspensions and drops (Figure 3).

**Figure 3.**
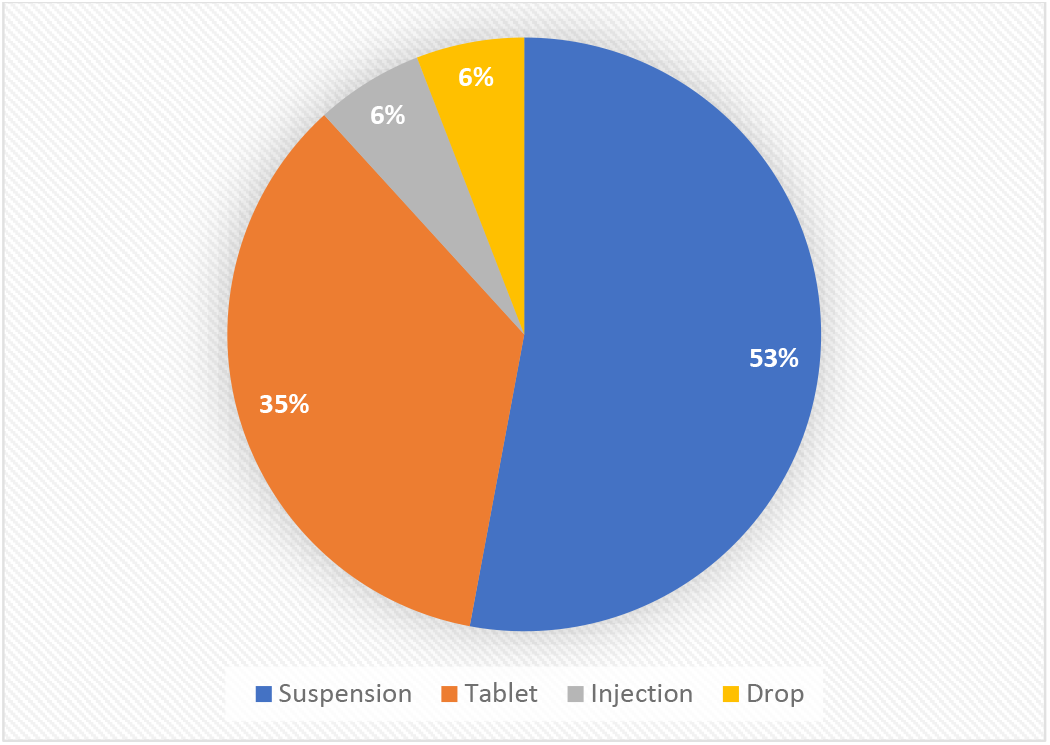
Prevalence of all three quality indicators - NAFDAC number, MAS, hologram - by dosage form type among 17 commercial products found to contain all three quality indicators in a sample of 386 commercial pharmaceutical products in a surveyed community pharmacy in Bayelsa State, Nigeria

The 29 products without a NAFDAC number also did not contain scratch codes, with all but 3 also not containing a hologram. Three of these were antimicrobial formulations comprising one parenteral antibiotic product with a hologram, an oral cotrimoxazole, and an eyedrop. These products were mostly (89%, 23/26) imports.

## Discussion

**Nigeria, in common with many LMICs face the double burden of high disease levels and high prevalence of SF medicines. This creates the need for continued vigilance and innovations to ensure the quality of medicines**. This study provides the first profiling of a comprehensive set of medicines packaging authentication strategies in Nigeria. It describes, using the inventory of a conveniently selected community pharmacy, three pre-defined quality assurance protocols on medicines packaging in Nigeria as a proxy risk ranking tool.

The NAFDAC number was the most prevalent authentication technology. Most products were registered by NAFDAC. Overall, less than 10% of the products were unregistered. This is in accordance with the general level of registered products in the market or supply chain in Nigeria, reported at 80-90% (11). In apparent conformity with NAFDAC’s implementation of the MAS, most antimicrobials had a scratch code. This finding of an almost 100% prevalence of MAS on antimicrobial packaging is in contrast to a study which found the prevalence of MAS to be between 60-79% in a sample of medicines outlets in Lagos (12). Holograms were the least prevalent authentication technology.

The presence of these authentication technologies does not certify quality. NAFDAC numbers, and holograms, have been known to be falsified (11,13,14). This threat has led to the development of a tool to check for fake NAFDAC numbers (15,16).

Medicines more prone to adulteration evince manufacturers’ ameliorative actions on the package. The increased risk of falsification of individual authentication technologies leads to the presence of multiple technologies, as concerned manufacturers make a greater investment to ensure product integrity as well as visually demonstrate this to the consumer.

Antimicrobials, as a therapeutic group, contained the most quality assurance markings on their packaging, in suggesting that they bear the greatest propensity, or risk, for being SF. Indeed, this is confirmed from the WHO study on SF medicines which found that antimalarials and anti-infective medicines were among four therapeutic groups most prone to failure, or to being substandard or falsified. The other two groups being anti-epileptics and genitourinary and sex hormone products (1).

The communicable disease burden in Nigeria is high. The rising trend in infections and the increasing spread of AMR make it necessary ensure the quality of antimicrobials in circulation in the country, and the need for continuous vigilance to guard against the entry of SF antimicrobials.

An interesting finding of this study is the relative high risk of SF in medicines for children.

Antibiotic medicines for children – suspensions and drops – bore the greatest number of authentication technologies. This data suggests the need to prioritize routine medicines quality assurance for antibiotic medicines for children. This conforms with reports indicating that children remain particularly susceptible to SF medicines (17–20)

There is the need, going by the presence of multiple quality assurance mechanisms on their packaging which presage risk, for greater vigilance during procurement of antimicrobials, as they are more likely to be SF, particularly for children medicines. Thus, in addition to this visual screening using packaging authentication technologies, a simple chemical quality assurance method would need to be employed at the facility level. The routine testing or assessment of high-risk products could help prevent pitfalls (avoid the negative health outcomes) associated with the use of SF medicines in contexts with non-stringent regulatory authorities.

Limitations: This analysis did not include all commercial pharmaceutical products that may be found in Nigeria. However, it covered representatives of all but two therapeutic groups as categorized by the WHO/ATC system. The results it presents may also not be generally representative for Nigeria, even though it is the first of such study. The analysis did not include registration numbers by foreign MRAs as these are usually not considered for quality assessments.

## Conclusion

Multiple modes of medicine authentication on packaging can predict SF risk. Antimicrobials, especially antibiotics for children, were the therapeutic group with the highest prevalence of authentication technologies or quality indicators on their packaging. **The implication of this finding is the need for health care facilities to implement strict medicines quality assurance procurement practices especially for antimicrobials. Equipping hospitals and community pharmacies with portable and easy-to-use devices to provide risk-based medicine quality assurance services is a much-needed public health intervention to help stem the tide of SF medicines in the global south. Consumer awareness programs should be elaborated and sustained**.

## Data Availability

All data produced in the present study are available upon reasonable request to the authors

## Acknowledgments

Mary David and Kelvin Leghemo for data collation.

